# Modulating Amygdala Responses in Major Depressive Disorder using Naturalistic Neurofeedback with Emotional Faces

**DOI:** 10.64898/2026.11.03.26367151

**Authors:** Cindy Sumaly Lor, Amelie Haugg, Martina Langer, David Willinger, Johanna Aubry, Kim Molinari, Philipp Stämpfli, Mengfan Zhang, Birgit Kleim, Annette Brühl, Ronald Sladky, Frank Scharnowski, Apurva Watve

## Abstract

Emotion regulation is critical to mental health, and its disruption can lead to psychiatric conditions such as major depressive disorder (MDD). MDD encompasses diverse symptoms, including anhedonia and anxiety, which may be associated with distinct patterns of altered neural activity. Interventions that address the neural mechanisms underlying these symptom dimensions may improve treatment efficacy. We investigated an adaptive, naturalistic real-time functional magnetic resonance imaging neurofeedback paradigm designed to differentially target anhedonia and anxiety through bidirectional amygdala regulation. Unlike conventional abstract feedback, our approach used emotional faces whose expressions changed dynamically according to ongoing amygdala activity, embedding neural self-regulation within a socially relevant emotional context. In this parallel-group study, 21 patients with MDD received one of two neurofeedback conditions. In the Happy-up group, patients attempted to increase amygdala activity by making a neutral face happier to enhance positive emotional responding. In the Fear-down group, patients attempted to decrease amygdala activity by making a fearful face less fearful to reduce negative emotional reactivity. Both groups showed the expected pattern of neural regulation during neurofeedback, with greater amygdala activity during Happy upregulation and decreasing amygdala activity during Fear downregulation. Following the intervention, patients showed reductions in depression severity, anxiety, and anhedonia, and more positive evaluations of neutral and happy faces. However, improvements in anhedonia and anxiety did not differ significantly between groups. These findings show bidirectional amygdala regulation through naturalistic neurofeedback in MDD. Although symptom-specific clinical effects were not established, the observed clinical improvements underscore the therapeutic potential of this approach.

## Introduction

Major depressive disorder (MDD) is a complex affective disorder associated with high rates of comorbidity and substantial global burden^1–4^. Although psychotherapy and pharmacotherapy approaches are effective for many patients, approximately one third do not respond adequately to first-line treatments^5^. Developing safe, non-invasive and non-pharmacological interventions that target clinically relevant neural mechanisms could therefore offer a valuable complement to existing treatments.

MDD is characterized by substantial clinical and neurobiological heterogeneity. Patients meeting the same diagnostic criteria can differ considerably in the type and severity of their symptoms, such as anhedonia, anxiety, dysphoria, cognitive impairment, and altered emotional reactivity^1,6^. This variability has motivated therapeutic approaches that focus on specific symptom dimensions and their underlying neurobiological substrates rather than assuming a uniform pathophysiology across MDD^1,3,7^. The Research Domain Criteria (RDoC) initiative, for instance, reoriented psychiatric research toward dimensional systems of functioning, including positive and negative valence processes, rather than categorical diagnoses^8^.

This distinction is particularly relevant to MDD, in which both positive and negative affective processing are altered. In particular, reduced responsiveness to positive emotional information has been linked to anhedonia^9,10^, whereas heightened or sustained responses to negative and threatening information may contribute to anxiety and dysphoria^11,12^. Both forms of altered affective processing involve the amygdala. This subcortical region contributes to the evaluation of emotional salience and to affective learning, and plays a particular role in processing facial expressions^13^. Consistent with this, studies and meta-analyses of MDD report alterations in amygdala responses in both directions, with blunted responses to positive information and heightened responses to negative stimuli^14–17^. These findings make the amygdala a plausible target for a neurofeedback intervention aimed at modulating affective processes relevant to anhedonia and anxiety.

Real-time functional magnetic resonance imaging neurofeedback (rtfMRI-NF) provides a non-invasive means of training voluntary regulation of spatially defined neural signals using feedback derived from ongoing brain activity^18–21^. Prior work has established rtfMRI-NF as a feasible method for modulating neural systems involved in emotion regulation in healthy participants^22,23^ and in patients with MDD^24–31^. Studies targeting the amygdala have demonstrated upregulation through positive autobiographical memory recall^26,27,32^ and downregulation during exposure to aversive stimuli in healthy participants^22,33–35^. Such approaches have been associated with improvements in depressive symptoms and emotional processing in patients with MDD^26,27,32,36,37^, as well as changes in prefrontal–limbic functional connectivity during downregulation in healthy participants^34^. Related work has shown that neurofeedback targeting prefrontal activity during cognitive reappraisal can also reduce amygdala responses to aversive stimuli^38^.

Previous rtfMRI-NF studies in MDD have largely relied on internally generated emotional or cognitive states, such as positive autobiographical memory recall or mental imagery, paired with abstract visual feedback displays^24–27^. The processing of social cues has thus received little attention in MDD neurofeedback research, although social functioning is a core aspect of MDD, with impairments that can persist beyond symptomatic remission^39,40^. Reduced responsiveness to social reward and difficulties interpreting others’ emotions and mental states may contribute to a cycle of social disengagement and diminished opportunities for positive interpersonal experiences^39,41^. Facial expressions are central to these interactions, yet MDD is associated with negatively biased interpretations and reduced recognition accuracy of positive, neutral, and ambiguous expressions^42–44^. Experimental training studies have further demonstrated that biases in the interpretation of ambiguous facial expressions can be modified^45^, while changes in facial-affect processing have been associated with improvements in depressive symptoms^43,46^. Socially meaningful stimuli such as emotional faces may therefore offer a complementary approach to neurofeedback in MDD. Quevedo et al.^47^ previously incorporated smiling self-faces into neurofeedback training in depressed adolescents, combining facial stimuli with positive autobiographical memory recall and abstract feedback. We previously developed a dynamic face-based amygdala neurofeedback paradigm and tested it in healthy participants^23^. In the present study, we adapted this paradigm for patients with MDD to investigate amygdala regulation in positive and negative social-affective contexts.

In this paradigm, the neurofeedback signal was conveyed dynamically through the face itself. Rather than representing ongoing amygdala activity through a separate visual display such as a bar or thermometer^19,35,48–50^, changes in amygdala activity were translated into gradual changes in the emotional intensity of the face stimulus (Fig. 1). The facial expression thus constituted both the affective context in which regulation occurred and the neurofeedback signal itself. This dynamic format may strengthen the affective impact of the feedback, as dynamic expressions, relative to static ones, can facilitate emotion perception, elicit facial mimicry, and evoke stronger affective and physiological responses^51^.

**Fig. 1.**
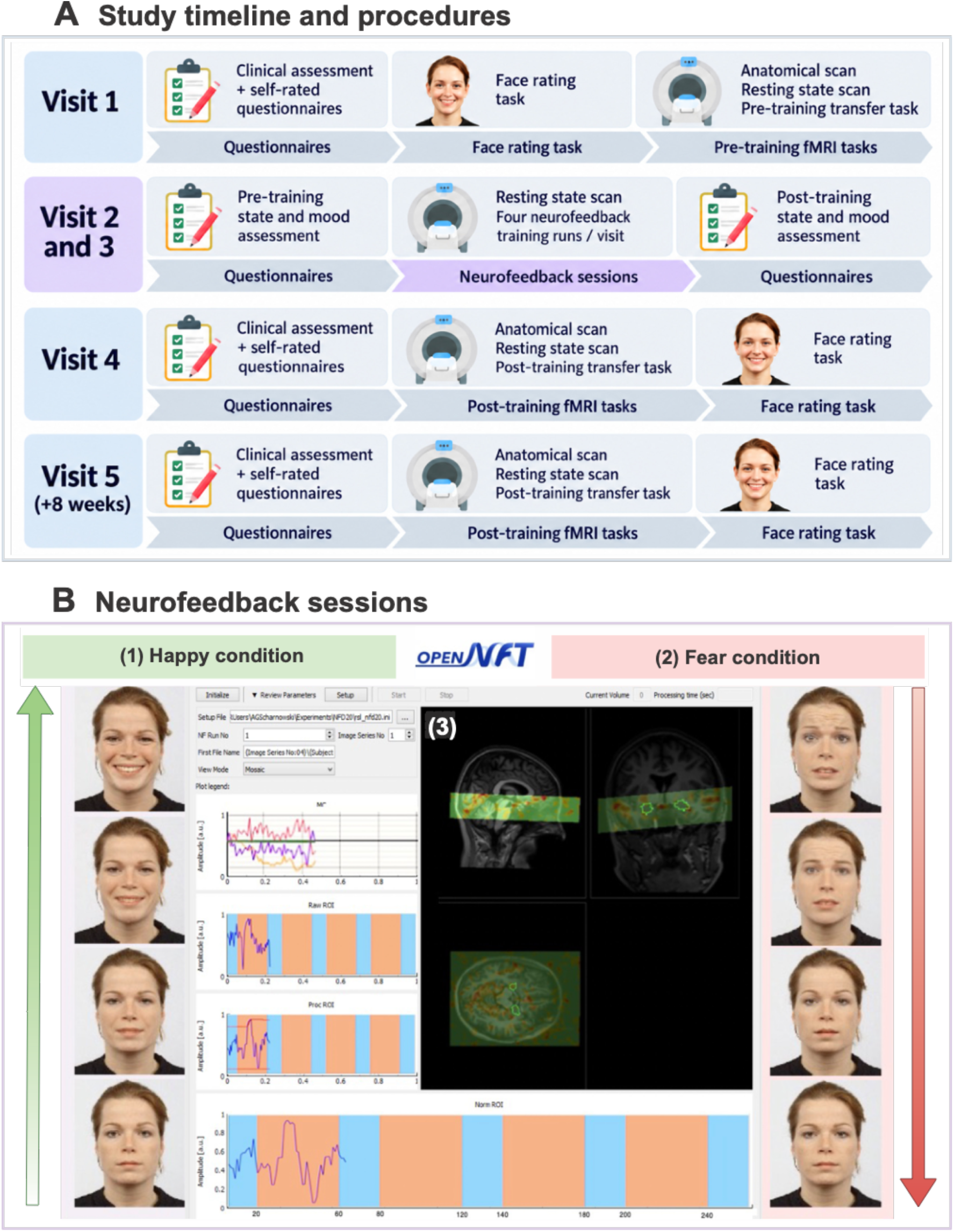
Study timeline and naturalistic emotional face neurofeedback paradigm. **(A)** Overview of the experimental protocol across five visits, including baseline assessments, two neurofeedback training sessions, post-training assessments, and an eight-week follow-up. (B) Overview of the adaptive face-based neurofeedback paradigm. (1) In the Happy-up condition, increases in bilateral amygdala activity shifted a neutral face toward a happier expression. (2) In the Fear-down condition, decreases in bilateral amygdala activity shifted a fearful face toward a more neutral expression. In both conditions, changes in amygdala activity dynamically modified facial expressions, with the resulting affective changes intended to further support regulation in the targeted direction. (3) The central panel shows the OpenNFT real-time fMRI neurofeedback interface, including the bilateral amygdala regions of interest used to derive the feedback signal and the raw and processed ROI time courses. Alternating background colors indicate baseline and regulation blocks.

The study paradigm included positive (Happy) and negative (Fear) emotion regulation conditions. In the Happy condition, successful amygdala upregulation caused a neutral face stimulus to become progressively happier. Positive facial feedback therefore depended on successful regulation, and this contingency was intended to reinforce amygdala upregulation and sustain engagement with positive emotional information. In the Fear condition, successful amygdala downregulation caused a fearful face to become gradually more neutral, reducing negative emotional input. In both conditions, the resulting change in affective input was intended to further shift amygdala activity towards the targeted regulatory direction, forming a self-reinforcing closed loop between amygdala activity and the emotional stimulus. Importantly, the feedback thereby became an active element of the regulatory loop rather than a passive display of brain activity.

Patients with MDD were assigned to either a Happy-up or Fear-down group. The Happy-up group received real neurofeedback for amygdala upregulation in the Happy condition, whereas the Fear-down group received real neurofeedback for amygdala downregulation in the Fear condition. Both groups additionally performed the opposite regulation task using yoked sham feedback, in which changes in facial expression were generated artificially based on the participant’s performance during real neurofeedback rather than their concurrent amygdala activity. Thus, both groups performed both regulation tasks but received real neurofeedback only for their assigned regulation direction.

This study addressed three main questions. First, we examined whether dynamic face-based amygdala neurofeedback was associated with clinical improvement, including changes in overall depression severity, anhedonia, anxiety, mood, and perceived sense of control in MDD patients. We hypothesized that depressive symptoms would improve across training, with greater reductions in anhedonia in the Happy-up group and greater reductions in anxiety in the Fear-down group.

Second, we tested whether patients could successfully regulate amygdala activity during neurofeedback and whether these effects generalized beyond the training context and persisted over time. We hypothesized that amygdala activity would be higher in the Happy condition with real feedback and lower in the Fear condition with real feedback than in the other regulation conditions, and that comparable regulation would be observed during a No-Feedback Transfer Task at post-training and follow-up. We additionally examined whether regulation changed across training runs.

Third, we examined whether neurofeedback affected emotional processing and subjective experience. We hypothesized that the Happy-up group would show greater changes in responses to positive emotional faces, whereas the Fear-down group would show greater changes in responses to negative faces. We further expected participants to report higher perceived regulation performance during real than sham feedback. Finally, in exploratory analyses, we examined whether individual differences in amygdala regulation were associated with clinical improvement.

## Methods

### Preregistration

The study protocol, intervention design, and outcome measures were registered in the ISRCTN trial registry (Ref. No. ISRCTN71745131). The registration specified depression severity as the primary clinical outcome and included measures of anhedonia, anxiety, mood, sense of control, emotional-face processing, and amygdala responses as secondary outcomes. The comparison of perceived regulation performance during real versus sham feedback was not preregistered and was therefore considered exploratory. Analyses examining associations between individual differences in amygdala regulation and clinical improvement were likewise exploratory.

### Participants

Twenty-two patients with a current episode of mild to moderate unipolar depression, diagnosed according to ICD-10 criteria (F32–F33), were enrolled in the study. Power calculations for the sample size are provided in the Supplementary Material. Patients who took antidepressants or oral contraceptives were required to be on a stable regimen. All patients were recruited by an experienced clinician according to established procedures at the Psychiatric University Hospital, Zurich, from internal depression wards and outpatient clinics. Recruited patients were between 18–65 years of age, fluent in German, right-handed, with normal vision, and without MRI contraindications such as pregnancy, brain surgery, or metal implants. Based on the Mini-International Neuropsychiatric Interview (MINI)^52^ and a semi-structured clinical interview, patients with Axis-I psychiatric disorders other than MDD (e.g., organic disorders, psychoactive substance use, psychotic and bipolar disorders, posttraumatic stress disorder, panic disorder, personality disorder, etc.) and other neurological comorbidities such as epilepsy, stroke, brain tumour, and traumatic brain injury were excluded from the study (N = 1). Eligible participants (N = 21) were alternately assigned to the Happy-up group (N = 10) and the Fear-down group (N = 11). Participants signed an informed consent form following the Declaration of Helsinki before participation. No compensation was offered for participation in the study. The Cantonal Ethics Committee of Zurich (Kantonale Ethikkommission Zürich), Switzerland, gave ethical approval for this work (BASEC No. 2017-01306). The study was conducted in accordance with the Declaration of Helsinki, and all participants provided written informed consent prior to participation.

### Study Overview

The study was conducted at the MR Centre of the Psychiatric University Hospital Zurich over four consecutive visits (V1–V4), with an additional follow-up visit (V5) scheduled 60 days after the first visit (Fig. 1). On the first visit (V1), the participants completed pre-training measurements, including a clinical assessment, a Face Rating Task, and a No-Feedback Transfer Task inside the MRI scanner. On the second (V2) and third (V3) visit, participants completed four neurofeedback (NF) training runs per day. On the fourth session (V4), participants completed a set of post-training assessments, which were identical to those administered at V1. The same assessments were repeated at the follow-up visit (V5), scheduled 60 days after V1, which corresponds to approximately eight weeks after the neurofeedback training. One participant from the Happy-up group completed the post-training clinical assessment and Face Rating Task at V4 but dropped out before the post-training MRI session and did not attend the follow-up visit (V5).

The current study followed the guidelines provided by the Consensus on Reporting and Experimental Design of Clinical and Cognitive-Behavioural Neurofeedback Studies (CRED-nf)^53^ and included the checklist summarizing the study design in the Supplementary Table 1.

### Region of Interest (ROI)

A high-resolution, probabilistic mask of the bilateral amygdala was derived from the California Institute of Technology CIT168 human brain atlas^54^. Using the normalization parameters estimated in SPM12 (Wellcome Centre for Human Neuroimaging, University College London, London, UK), this atlas-based ROI was nonlinearly transformed from Montreal Neurological Institute (MNI) space into each participant’s native anatomical space by inverse warping (Fig. 1B).

### Face Stimuli

Thirty gender-balanced faces of Caucasian models depicting neutral, fearful, and happy emotions were selected from the Radboud Face Database^55^. The No-Feedback Transfer Task and the Face Rating Task used the original static face stimuli. For the neurofeedback training, dynamic emotional faces were generated using a Python-based face morphing algorithm^56^. Each face was morphed along a continuum from a neutral expression to either a happy or fearful expression, depending on the regulation condition. The resulting morphs enabled smooth, video-like transitions between emotional expressions, with Morph level 0 corresponding to a fully neutral expression and level 30 to the maximum emotional intensity of the happy or fearful expression. During neurofeedback, the displayed emotional intensity was adjusted in real time according to the mean blood-oxygen-level-dependent (BOLD) signal extracted from the participant’s bilateral amygdala.

### Naturalistic Face Feedback

Neurofeedback signals were estimated and presented using Open NeuroFeedback Training (OpenNFT), an open-source neurofeedback framework implemented in Python and MATLAB^57^. The feedback was scaled to the normalized amygdala time-series using OpenNFT’s default dynamic range to estimate the maximum and minimum limits of the scaling by using the average of the 5% highest and lowest signal intensities observed so far. The preprocessed amygdala signal was continuously mapped onto the emotional intensity of the face stimulus (morph levels 0–30), such that changes in amygdala activity produced smooth, dynamic transitions in facial expression rather than discrete changes between static images.

### Task Description

#### Neurofeedback Task

Participants completed eight NF runs spread over two training sessions (four runs per session). A NF training run consisted of four 40-s regulation blocks, interleaved with four 20-s baseline blocks in which participants mentally counted digits backwards while fixating a dot.

Each run included two Happy condition blocks and two Fear condition blocks, presented in alternating order. During the Happy condition, participants viewed a dynamic face varying along a neutral-to-happy continuum and were instructed to “make the face happier.” During the Fear condition, they viewed a face varying along a neutral-to-fearful continuum and were instructed to “make the fearful face neutral.” Facial expression intensity was updated once per second, and a different face identity was used in each block to reduce habituation (Fig. 2). Before training, participants were informed about the amygdala’s role in emotional processing and that changes in the facial expressions reflected their ongoing amygdala activity. Participants were free to select their own emotion-regulation strategies and were asked to verbalize their intended strategies before training. They were instructed to remain focused on the regulation task throughout each run and could adapt or change their strategies between runs.

**Fig. 2.**
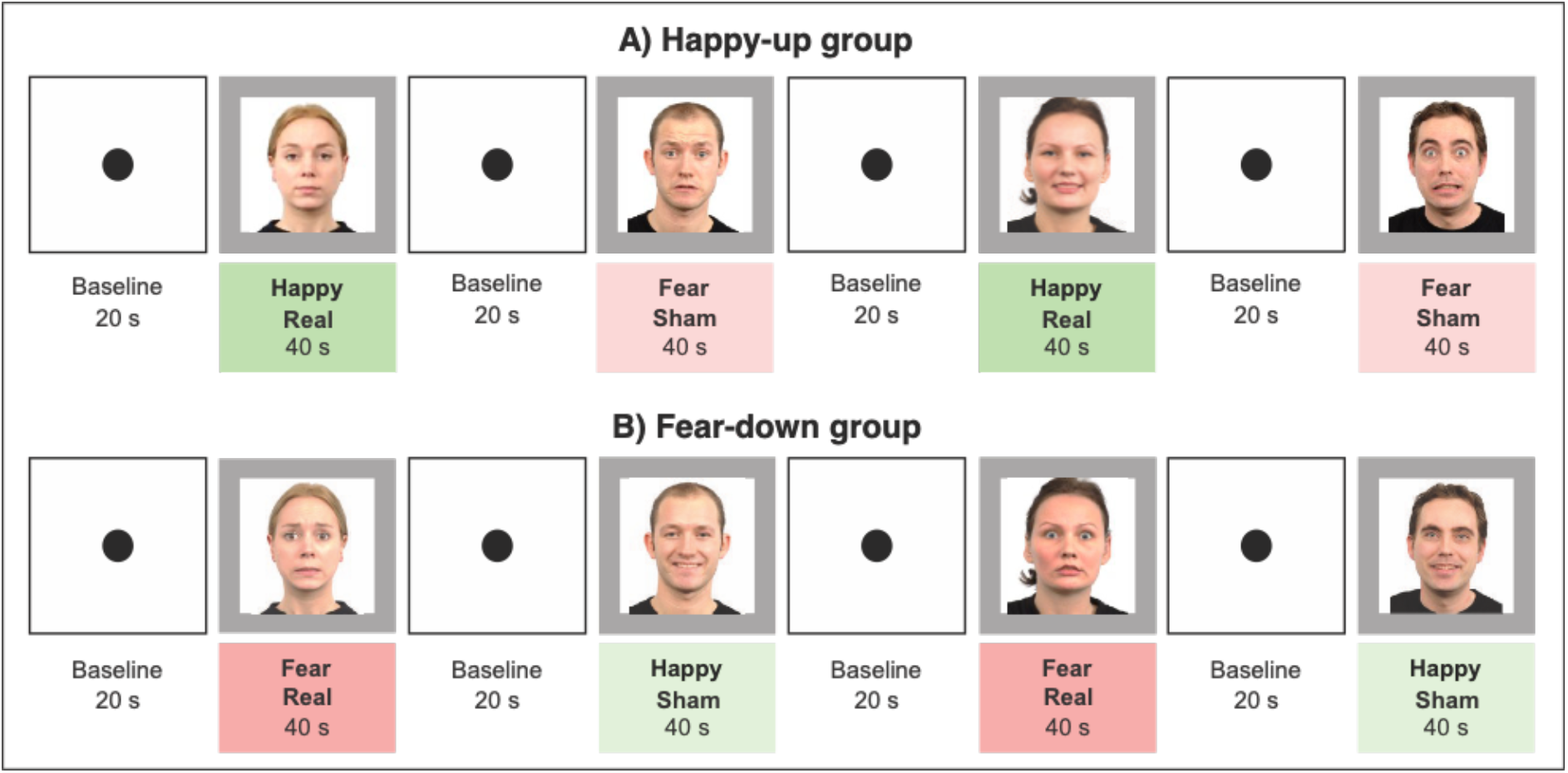
Neurofeedback task design. Each neurofeedback run comprised four 40-s regulation blocks interleaved with four 20-s baseline blocks. During baseline blocks, participants counted backward while fixating on a dot. The Happy-up group received real neurofeedback during Happy upregulation and yoked sham feedback during Fear downregulation, whereas the Fear-down group received real neurofeedback during Fear downregulation and yoked sham feedback during Happy upregulation. During real feedback, the emotional expression of the face was continuously updated according to ongoing bilateral amygdala activity.

The regulation blocks provided either real or yoked sham feedback to the participant, which allowed for a within-subject control comparison^58^. During real feedback, the participant’s ongoing amygdala activity directly controlled the emotional intensity of the face. In the Happy condition with real feedback (Happy Real), increases in amygdala activity shifted the face toward a happier expression. In the Fear condition with real feedback (Fear Real), decreases in amygdala activity shifted the face toward a more neutral expression. In both conditions, the system was designed to establish an adaptive, self-reinforcing closed loop between amygdala activity and the emotional expression of the face stimulus. Changes in amygdala activity dynamically altered the facial expression, which in turn was intended to reinforce amygdala regulation towards the targeted direction.

The participants were assigned to either of two groups. The Happy-up group received real feedback during the Happy condition only (Happy Real), and sham feedback during the Fear condition (Fear Sham) (Fig. 2A). The Fear-down group received real feedback during the Fear condition only (Fear Real), and sham feedback during the Happy condition (Happy Sham) (Fig. 2B). Importantly, this double-dissociation design allowed both groups to complete both the Happy and Fear conditions while receiving contingent neurofeedback in only the condition corresponding to their assigned training group.

At the end of each NF training run, participants rated their perceived performance in Happy and Fear conditions on a scale from 1 (= bad) to 10 (= good). Mean performance ratings were used to compare perceived regulation success during real and sham feedback.

### No-Feedback Transfer Task

Participants completed the No-Feedback Transfer Task at pre-training, post-training, and follow-up (V1, V4, and V5). The task followed the same block structure as the neurofeedback task, with four 40-s regulation blocks (without feedback) interleaved with four 20-s baseline blocks. Each regulation block comprised five static face trials presented for 8 s each.

During the Fear condition, participants viewed fearful faces and applied emotion regulation strategies to downregulate amygdala activity. During the Neutral condition, participants viewed neutral faces and applied positive regulation strategies with the aim of upregulating amygdala activity. Importantly, no neurofeedback was provided during this task, and the facial expressions therefore remained static rather than adapting as a function of ongoing amygdala activity.

### Face Rating Task

Participants’ perceptual responses (affective perception) and arousal levels (affective arousal) in response to emotional expressions were assessed using a face rating task outside the MRI scanner at all three assessment timepoints. The task consisted of 60 gender-balanced face trials (20 trials/emotion), which were presented in a pseudo-randomized fashion using PsychoPy (version 3.1) software^59^. Each trial began with a fixation dot presented for 1 s, followed by the emotional face presented for 3 s. The faces were either happy, neutral, or fearful. Participants were then asked, “What do you think this person is feeling?” (German: “Was denken Sie fühlt diese Person?”), which was used as a measure of affective perception, and “How do you feel when you look at this face?” (German: “Wie fühlen Sie sich bei der Betrachtung dieses Gesichts?”), which was used as a measure of affective arousal. Participants answered these questions by clicking on a scale from 0 to 100, with the lowest and highest anchors corresponding to ‘anxious’ and ‘happy’ (German: ‘ängstlich’ and ‘glücklich’) for affective perception, and ‘relaxed’ and ‘excited’ (German: ‘entspannt’ and ‘aufgeregt’) for affective arousal. No time limit was specified to answer these questions. The task took approximately ten minutes to complete.

### Brain Image Acquisition

Brain scans were acquired with an Achieva 3-Tesla MRI scanner (Philips Healthcare, Best, The Netherlands) using the manufacturer’s 32-channel head coil at the MR Centre of the Psychiatric University Hospital Zurich. At the beginning of the scanning session, a whole-brain T1-weighted structural scan (TR/TE = 9.3/4.6 ms, 160 sagittal slices, reconstructed voxel size = 1 × 1 × 1 mm^3^, field of view = 240 × 240 × 160 mm^3^, flip angle = 8°, acquisition time = 3 min 44 s) was acquired for anatomical reference.

A single whole-brain echo-planar imaging (EPI) volume was then acquired using the same EPI contrast and in-plane resolution as the subsequent neurofeedback acquisition, but with extended slice coverage, for coregistration of the partial-volume neurofeedback images to the T1-weighted anatomical image (TR/TE = 6000/35 ms, 70 axial slices, voxel size = 2 × 2.04 × 2 mm^3^, matrix size = 112 × 110, field of view = 224 × 224 × 140 mm^3^, flip angle = 65°, SENSE factor = 2, acquisition time = 42 s).

For each neurofeedback run, 215 volumes were acquired using a partial-volume gradient-echo T2*-weighted EPI sequence restricted to the target regions, including the medial orbitofrontal cortex, temporal lobe, amygdala, fusiform gyrus, and parts of the visual cortex (TR/TE = 1250/35 ms, 16 axial slices, voxel size = 2 × 2.04 × 2 mm^3^, slice gap = 1 mm, matrix size = 112 × 110, field of view = 224 × 224 × 47 mm^3^, flip angle = 65°, SENSE factor = 2, acquisition time = 4 min 33 s per run). The first five acquired volumes were excluded from subsequent analysis.

Neural data were exported in real time to a high-performance computer using the proprietary Philips DRIN export system. Feedback from the bilateral amygdala was presented continuously using OpenNFT^57^.

### Clinical Assessment

Clinical changes were evaluated using standardized psychometric questionnaires at pre-training, post-training, and follow-up. Depression severity was assessed using the 21-item Hamilton Depression Rating Scale (HAM-D)^60^ and the Montgomery–Åsberg Depression Rating Scale (MADRS)^61^ as primary outcome measures. Anxiety was assessed using the State–Trait Anxiety Inventory (STAI)^62^ and anhedonia using the German Snaith–Hamilton Pleasure Scale (SHAPS)^63^, summed across 14 items. Mood was assessed using the 12-item short form of the Multidimensional Mood State Questionnaire (German: MDBF)^64^, with item responses summed after reverse scoring to yield good-mood (MDBF-GS), wakefulness (MDBF-WM), and calmness (MDBF-RU) subscale scores. Competence and control beliefs were assessed using the FKK-SKI-PC composite of the Competence and Control Beliefs Questionnaire (German: FKK)^65^, expressed as standardized T-scores (normative mean = 50, SD = 10), hereafter referred to as “sense of control.” Higher scores on the HAM-D, MADRS, STAI, and SHAPS indicate greater symptom severity, whereas higher scores on the MDBF subscales and FKK-SKI-PC indicate better mood, greater wakefulness and calmness, and stronger internal relative to external control beliefs, respectively.

### Data Analysis

#### Online preprocessing and neurofeedback signal calculation

An SPM12-based spatial transformation pipeline was implemented to align each participant’s structural scan, whole-brain EPI volume, test EPI volume, and a bilateral amygdala mask into the subject’s space that served as a volume of interest during NF training. Online fMRI data preprocessing and feedback signal calculation were performed using OpenNFT’s default pipeline which includes realignment for motion correction and spatial smoothing with a 5 mm full width at half maximum (FWHM) Gaussian kernel. Spike removal using a Kalman filter and drift removal using a cumulative General Linear Model (GLM) were performed to process temporal NF data^66^. The resulting preprocessed amygdala time series were extracted and used as a neurofeedback signal.

#### Offline Brain Data Analysis

Offline data processing was performed with SPM12 and included slice-timing correction, realignment, coregistration, normalization to MNI space, and spatial smoothing with a 6 mm FWHM Gaussian kernel.

Separate first-level GLMs were fitted for each neurofeedback run, with two regressors of interest for the real- and sham-feedback regulation blocks. The real-feedback regressor represented the Happy condition in the Happy-up group and the Fear condition in the Fear-down group, with two 40-s blocks beginning at 20 and 140 s. The sham-feedback regressor represented the opposite condition, with two 40-s blocks beginning at 80 and 200 s.

Separate GLMs were also fitted for the pre-training, post-training and follow-up runs, with regressors for the Fear and Neutral conditions. Block durations and onsets were identical to those of the neurofeedback runs. In the Fear-down group, fear blocks began at 20 and 140 s and neutral blocks at 80 and 200 s; this order was reversed in the Happy-up group. All task regressors were convolved with SPM’s canonical hemodynamic response function.

All models included six realignment parameters and the mean white-matter signal as nuisance regressors. Volumes with framewise displacement (FD) above 0.5 mm were censored via spike regression, with a separate nuisance regressor for each affected volume. A 128-s high-pass filter was applied, and temporal autocorrelation was modelled with an AR(1) process. Models were estimated with SPM12’s classical estimation procedure.

Runs were excluded from the analysis if more than 20% of volumes were censored or head displacement exceeded 3 mm in any direction.

## Statistical Analysis

### 1. Clinical and symptom-specific effects

The primary outcome of interest was the change in depression severity from pre- to post-training, assessed with HAM-D and MADRS scores. Separate linear mixed-effects models (LMMs) were fitted in R using the lme4 and lmerTest packages with restricted maximum likelihood estimation. Each model included time (pre-training, post-training, follow-up), group (Happy-up, Fear-down) and their interaction as fixed effects, with a random intercept for participants. Time was dummy-coded with pre-training as the reference, and group was effect-coded (Happy-up = +0.5, Fear-down = −0.5). Time coefficients therefore represent changes from pre-training averaged across the two groups, and interaction coefficients represent between-group differences in change (Happy-up minus Fear-down). The pre-training to follow-up contrast assessed longer-term changes. All available data were included (n = 21); one participant from the Happy-up group did not complete the follow-up assessment and contributed pre- and post-training data only.

Equivalent LMMs assessed changes in anhedonia (SHAPS), trait and state anxiety (STAI-T and STAI-S), mood (MDBF good mood, wakefulness and calmness) and sense of control (FKK-SKI-PC). Holm– Bonferroni correction was applied separately to the pre–post contrasts for anhedonia and anxiety (three tests) and for mood and sense of control (four tests). HAM-D and MADRS p-values were reported without adjustment. Time × group interactions tested the hypothesized symptom-specific effects; these and the follow-up contrasts are reported with unadjusted p-values. Effect estimates are presented as unstandardized regression coefficients (b) with 95% confidence intervals, and two-sided p-values were obtained from t-tests with Satterthwaite-approximated degrees of freedom.

### 2. Neural regulation and transfer

#### 2a. Neurofeedback regulation performance and improvement

Neurofeedback performance and neurofeedback improvement were analysed separately, following the terminology of Haugg et al.^67^. Neurofeedback performance referred to the extent to which amygdala activity differed between the four regulation conditions in the predicted direction, whereas neurofeedback improvement referred to changes in regulation performance over the course of training. All neural LMMs were fitted in R using the lme4 and lmerTest packages with restricted maximum likelihood estimation, and degrees of freedom for all tests were estimated using the Satterthwaite approximation.

Mean amygdala beta values were extracted for each run and condition from the SPM first-level beta maps. Runs with excessive head motion were excluded (18 runs), leaving 300 observations from 21 participants. Condition was modelled as a four-level categorical factor comprising Happy Real, Happy Sham, Fear Real and Fear Sham. Participants in the Happy-up group completed the Happy Real and Fear Sham conditions, and participants in the Fear-down group completed the Fear Real and Happy Sham conditions; comparisons between conditions completed by different groups were therefore between-subjects. Run was entered as a continuous, mean-centred predictor.

For neurofeedback performance, beta values were analysed using an LMM with condition and run as fixed effects and a random intercept for participants. We tested two hypotheses: Happy Real should elicit higher amygdala activity than each of the other three conditions, and Fear Real should elicit lower amygdala activity than each of the other three conditions.

For neurofeedback improvement, condition-specific linear changes across the eight neurofeedback runs were estimated using an LMM with condition, run and their interaction as fixed effects and a random intercept for participants, allowing a separate run slope for each condition. We tested two hypotheses: the Happy Real slope should be greater than the slope of each of the other three conditions, and the Fear Real slope should be lower than the slope of each of the other three conditions. Each condition’s slope was additionally tested against zero (one-sided in the predicted direction for the real conditions, two-sided for the sham conditions).

Each prediction was tested with three one-sided contrasts comparing the target condition with each of the other three conditions, using Satterthwaite-approximated degrees of freedom. A prediction was considered supported when the three contrasts were significant.

#### 2b. Transfer of amygdala regulation

Transfer effects of NF training were assessed in the No-Feedback Transfer Task by comparing changes in amygdala activity from the pre-training baseline to the post-training transfer assessment and the follow-up. Mean amygdala beta values were extracted for the Fear and Neutral conditions, and the Fear − Neutral difference was computed for each participant and session. An LMM was fitted with time (baseline, post-training, follow-up; baseline as reference), group (Happy-up, Fear-down; effect-coded as +0.5/−0.5) and their interaction as fixed effects, and a random intercept for participants. Time coefficients represent changes from baseline averaged across groups, and interaction coefficients represent between-group differences in change (Happy-up minus Fear-down). Sessions excluded for excessive head motion (n = 2) were omitted, and all remaining available data were included (21 participants, 59 sessions). Effect estimates are reported as unstandardized regression coefficients (b) with 95% confidence intervals, and p-values were obtained from t-tests with Satterthwaite-approximated degrees of freedom.

### 3. Emotional perception and subjective performance

#### 3a. Emotional perception and arousal

Behavioural changes in affective perception (or valence) and affective arousal were assessed based on the participant’s subjective ratings of emotional face stimuli in the Face Rating Task. An LMM analysis was performed for each emotion (neutral, fear, and happy), with affective perception and affective arousal ratings as dependent variables, time (pre-, post-training, and follow-up), group (Happy-up and Fear-down) and their interaction as fixed effect factors with a random intercept for each participant. Time was dummy-coded with pre-training as the reference, and group was effect-coded (Happy-up = +0.5, Fear-down = −0.5).

#### 3b. Subjective neurofeedback performance

Subjective self-regulation performance was assessed using participants’ ratings of their perceived regulation success after each NF run. Ratings were analysed using an LMM with feedback (real, sham), emotion (happy, fear), run, and their interactions as fixed effects and a random intercept for participants. Feedback and emotion were sum-coded, and run was entered as a continuous, mean-centred predictor. A directional contrast compared ratings in the real-feedback conditions with ratings in the sham-feedback conditions. Subjective improvement was assessed with the Feedback × Run interaction and by estimating condition-specific linear changes in ratings across runs. Positive slopes were expected for both real-feedback conditions, reflecting increasing perceived regulation success over training.

All behavioural LMMs were fitted in R using the lme4 and lmerTest packages with restricted maximum likelihood estimation, and p-values were obtained from t-tests with Satterthwaite-approximated degrees of freedom.

### 4. Brain–clinical associations

Associations between amygdala activity during NF training and clinical change were examined separately for each group. For each participant, amygdala beta values were averaged across all motion-eligible runs of the group-specific real-feedback condition: Fear Real for the Fear-down group and Happy Real for the Happy-up group. Clinical change scores were calculated as post-training minus pre-training scores for HAM-D, MADRS, STAI-T, STAI-S, SHAPS, FKK-SKI-PC, and the three MDBF subscales. For each clinical measure, a Spearman partial correlation tested the association between mean amygdala activity and the clinical change score, controlling for the pre-training score of the same measure. As a control analysis, the same correlations were computed for the group-specific sham condition. Two-sided p-values were adjusted within each group and condition across the nine clinical outcomes using the Holm–Bonferroni procedure.

## Results

### Sample characteristics

Participants were 21 patients with MDD, of whom 10 were assigned to the Happy-up group and 11 to the Fear-down group. Baseline demographic and clinical characteristics are summarized in Table 1. The groups did not differ significantly in sex distribution or in any baseline clinical measure (all p > .16), with comparable levels of depression severity, anhedonia, trait anxiety, and sense of control.

**Table 1.**
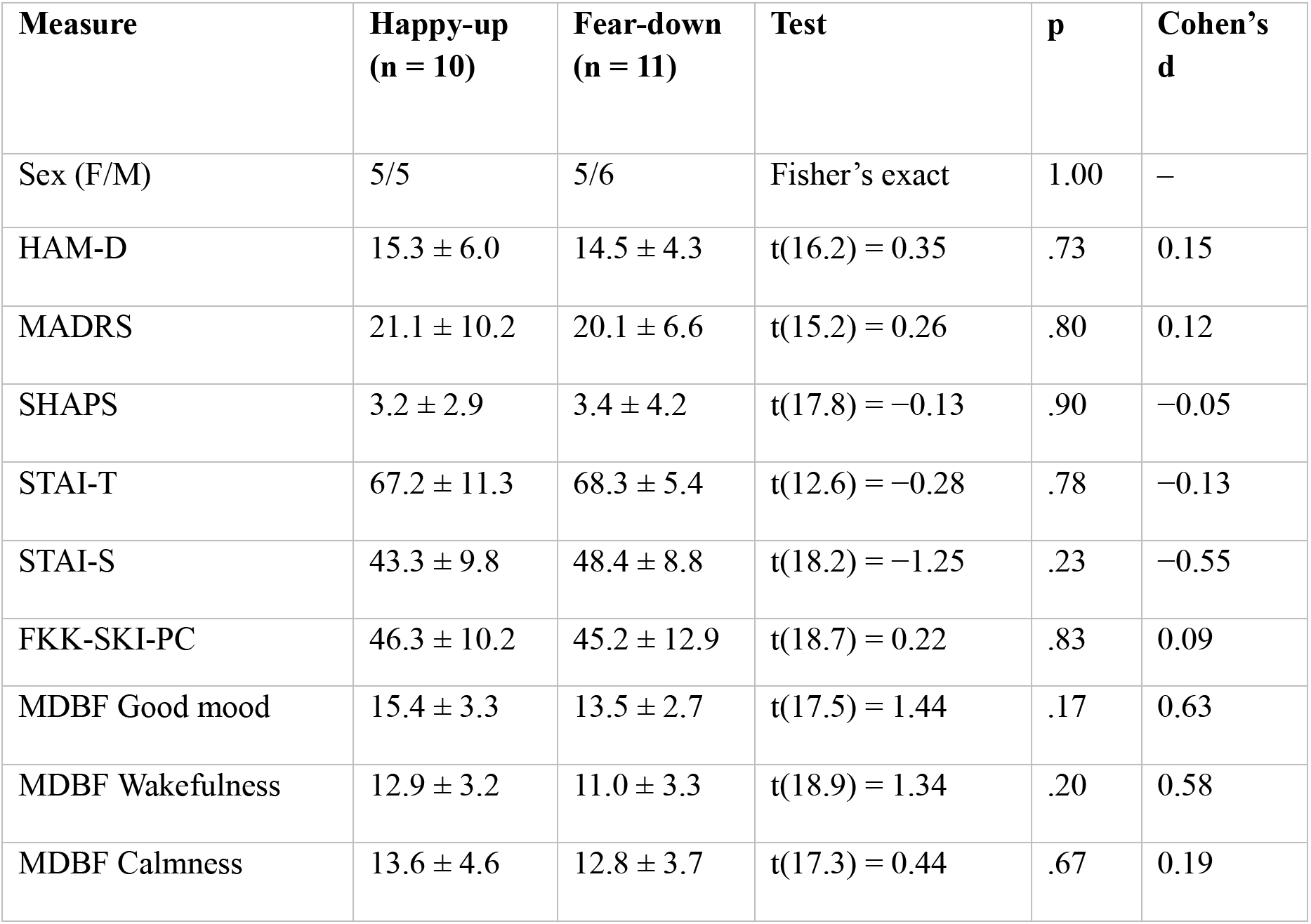
Baseline demographic and clinical characteristics of participants by neurofeedback group. *Note:* Values are mean ± SD unless otherwise indicated. Continuous variables were compared using Welch’s t-test (unequal variances); sex was compared using Fisher’s exact test. No baseline differences reached significance (all p > .16).

| **Measure** | **Happy-up (n = 10)** | **Fear-down (n = 11)** | **Test** | **p** | **Cohen’s d** |
|---|---|---|---|---|---|
| Sex (F/M) | 5/5 | 5/6 | Fisher’s exact | 1.00 | – |
| HAM-D | 15.3 ± 6.0 | 14.5 ± 4.3 | t(16.2) = 0.35 | .73 | 0.15 |
| MADRS | 21.1 ± 10.2 | 20.1 ± 6.6 | t(15.2) = 0.26 | .80 | 0.12 |
| SHAPS | 3.2 ± 2.9 | 3.4 ± 4.2 | t(17.8) = −0.13 | .90 | −0.05 |
| STAI-T | 67.2 ± 11.3 | 68.3 ± 5.4 | t(12.6) = −0.28 | .78 | −0.13 |
| STAI-S | 43.3 ± 9.8 | 48.4 ± 8.8 | t(18.2) = −1.25 | .23 | −0.55 |
| FKK-SKI-PC | 46.3 ± 10.2 | 45.2 ± 12.9 | t(18.7) = 0.22 | .83 | 0.09 |
| MDBF Good mood | 15.4 ± 3.3 | 13.5 ± 2.7 | t(17.5) = 1.44 | .17 | 0.63 |
| MDBF Wakefulness | 12.9 ± 3.2 | 11.0 ± 3.3 | t(18.9) = 1.34 | .20 | 0.58 |
| MDBF Calmness | 13.6 ± 4.6 | 12.8 ± 3.7 | t(17.3) = 0.44 | .67 | 0.19 |

### 1. Clinical and symptom-specific effects

#### 1a. Overall depression severity

Depression severity decreased following neurofeedback training. Mean HAM-D scores decreased from 14.90 (SD = 5.06) at pre-training to 10.86 (SD = 4.07) at post-training and were 11.90 (SD = 4.44) at follow-up. Relative to pre-training, HAM-D scores were significantly lower at post-training, b = −4.07, SE = 1.14, p = .001, 95% CI [−6.38, −1.75], and remained significantly lower at follow-up, b = −3.11, SE = 1.16, p = .011, 95% CI [−5.47, −0.75] (Fig. 3A).

**Fig. 3.**
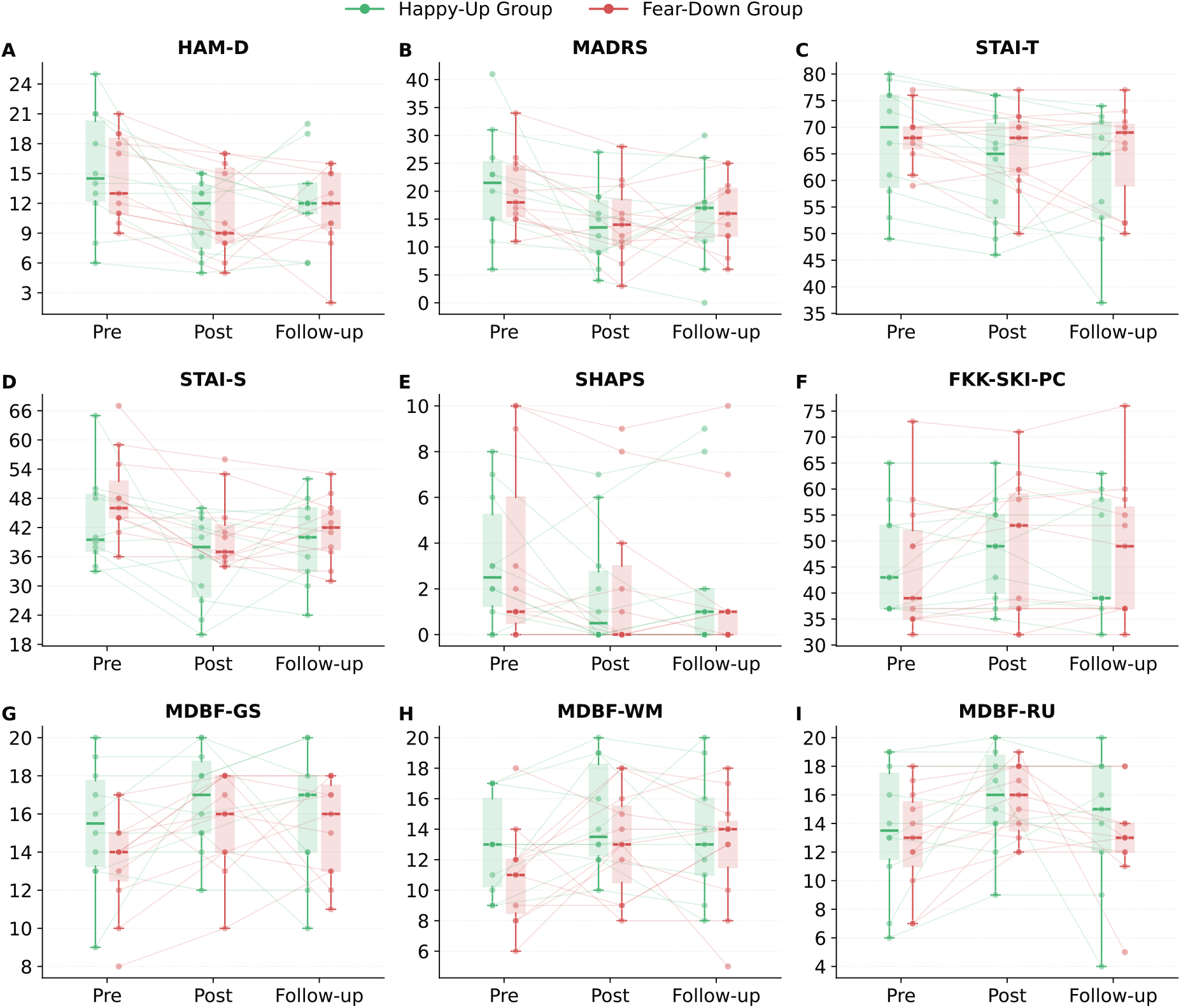
Changes in clinical and self-reported outcomes across the intervention. Scores are shown at pre-training, post-training, and follow-up separately for the Happy-up and Fear-down groups. Individual trajectories and distributions are shown for (A) HAM-D, (B) MADRS, (C) STAI-T, (D) STAI-S, (E) SHAPS, (F) FKK-SKI-PC, (G) MDBF good mood, (H) MDBF wakefulness, and (I) MDBF calmness. Depression severity, anhedonia, and anxiety improved over time, whereas improvements in sense of control and mood did not survive correction for multiple comparisons. No significant time × group interactions were observed.

Mean MADRS scores decreased from 20.57 (SD = 8.27) at pre-training to 14.05 (SD = 6.91) at post-training and were 16.10 (SD = 7.63) at follow-up. Relative to pre-training, MADRS scores were significantly lower at post-training, b = −6.57, SE = 2.02, p = .002, 95% CI [−10.66, −2.48], and remained significantly lower at follow-up, b = −4.81, SE = 2.05, p = .025, 95% CI [−8.97, −0.65] (Fig. 3B).

The time × group interactions were not significant for either measure. For HAM-D, the interaction was not significant from pre-to post-training, b = −0.86, SE = 2.29, p = .708, 95% CI [−5.50, 3.77], or from pre-training to follow-up, b = −0.04, SE = 2.33, p = .988, 95% CI [−4.75, 4.68]. Similarly, the MADRS interaction was not significant from pre-to post-training, b = −1.86, SE = 4.04, p = .647, 95% CI [−10.05, 6.32], or from pre-training to follow-up, b = −1.98, SE = 4.11, p = .632, 95% CI [−10.31, 6.34]. Thus, there was no evidence that changes in overall depression severity differed between the Happy-up and Fear-down groups.

#### 1b. Anhedonia and anxiety

Anxiety significantly decreased over time. Mean STAI-T scores decreased from 67.76 (SD = 8.51) at pre-training to 64.05 (SD = 9.39) at post-training and were 63.10 (SD = 10.87) at follow-up (Fig. 3C). Relative to pre-training, STAI-T scores were significantly lower at post-training, b = −3.76, SE = 1.80, p_adj_ = .043, p = .043, 95% CI [−7.41, −0.12], and at follow-up, b = −5.14, SE = 1.83, p = .008, 95% CI [−8.86, −1.43].

Mean STAI-S scores decreased from 45.95 (SD = 9.40) at pre-training to 38.05 (SD = 8.82) at post-training and were 40.55 (SD = 7.76) at follow-up (Fig. 3D). State anxiety was significantly lower than at pre-training both at post-training, b = −7.91, SE = 2.36, p_adj_ = .006, p = .002, 95% CI [−12.69, −3.12], and at follow-up, b = −5.51, SE = 2.40, p = .028, 95% CI [−10.37, −0.64].

Anhedonia decreased following neurofeedback training. Mean SHAPS scores decreased from 3.29 (SD = 3.51) at pre-training to 2.05 (SD = 2.97) at post-training and were 2.10 (SD = 3.37) at follow-up (Fig. 3E). Relative to pre-training, SHAPS scores were significantly lower at post-training, b = −1.24, SE = 0.51, p_adj_ = .041, p = .021, 95% CI [−2.28, −0.20], and remained significantly lower at follow-up, b = −1.24, SE = 0.52, p = .024, 95% CI [−2.30, −0.17].

No significant time × group interactions were observed for SHAPS, STAI-T, or STAI-S, providing no evidence that changes in anhedonia or anxiety differed between the Happy-up and Fear-down groups.

#### 1c. Mood and sense of control

Perceived sense of control increased numerically following NF training (Fig. 3F). Mean FKK-SKI-PC scores increased from 45.71 (SD = 11.41) at pre-training to 48.57 (SD = 11.75) at post-training and were 47.65 (SD = 12.52) at follow-up. Relative to pre-training, scores were higher at post-training, b = 2.83, SE = 1.30, p = .036, 95% CI [0.19, 5.46], but this effect did not survive correction for multiple comparisons, p_adj_ = .052. The increase at follow-up did not reach significance, b = 2.31, SE = 1.33, p = .090, 95% CI [−0.38, 5.01].

Mood also improved numerically after training, although none of the effects survived correction for multiple comparisons (Fig. 3G). Mean MDBF-GS good-mood scores increased from 14.43 (SD = 3.09) at pre-training to 16.14 (SD = 2.65) at post-training and were 15.50 (SD = 2.96) at follow-up. Good mood was higher at post-training, b = 1.70, SE = 0.71, p = .022, p_adj_ = .052, 95% CI [0.26, 3.13], whereas the increase at follow-up did not reach significance, b = 1.14, SE = 0.72, p = .121, 95% CI [−0.32, 2.60].

Mean MDBF-WM wakefulness scores (Fig. 3H) increased from 11.90 (SD = 3.32) at pre-training to 13.95 (SD = 3.50) at post-training and were 13.15 (SD = 3.88) at follow-up. Wakefulness was higher at post-training, b = 2.04, SE = 0.80, p = .015, p_adj_ = .052, 95% CI [0.43, 3.66], but the increase at follow-up did not reach significance, b = 1.33, SE = 0.81, p = .109, 95% CI [−0.31, 2.98].

Mean MDBF-RU calmness scores increased from 13.19 (SD = 4.08) at pre-training to 15.71 (SD = 3.05) at post-training and were 13.45 (SD = 4.16) at follow-up (Fig. 3I). Calmness was higher at post-training, b = 2.51, SE = 0.96, p = .013, p_adj_ = .052, 95% CI [0.56, 4.46], but not at follow-up, b = 0.42, SE = 0.98, p = .673, 95% CI [−1.57, 2.40].

None of the time × group interactions for control beliefs or mood were significant, providing no evidence that these changes differed between the Happy-up and Fear-down groups.

Overall, depression severity, anhedonia, and anxiety decreased following training, with these effects persisting at follow-up, while sense of control and mood showed nominal improvements at post-training that did not survive correction for multiple comparisons. These findings support our primary hypothesis of reduced depressive symptoms following neurofeedback training. However, none of these changes differed significantly between the Happy-up and Fear-down groups, providing no evidence for the hypothesized symptom-specific effects.

### 2. Neural regulation and transfer

#### 2a. Neurofeedback regulation performance and improvement

The planned LMM contrasts supported several of the predicted amygdala responses during neurofeedback (Fig. 4A). Amygdala activity was significantly higher in the Happy Real condition than in both the Happy Sham condition, b = 0.53, SE = 0.27, t(18.1) = 2.00, p = .031, and the Fear Sham condition, b = 0.31, SE = 0.10, t(272.3) = 3.08, p = .001, as well as the Fear Real condition, b = 0.58, SE = 0.27, t(18.1) = 2.19, p = .021. The differences between Fear Real and Happy Sham, b = 0.05, SE = 0.09, t(272.3) = 0.56, p = .287, and between Fear Real and Fear Sham, b = 0.28, SE = 0.27, t(18.1) = 1.04, p = .156, were in the predicted direction but did not reach significance.

**Fig. 4.**
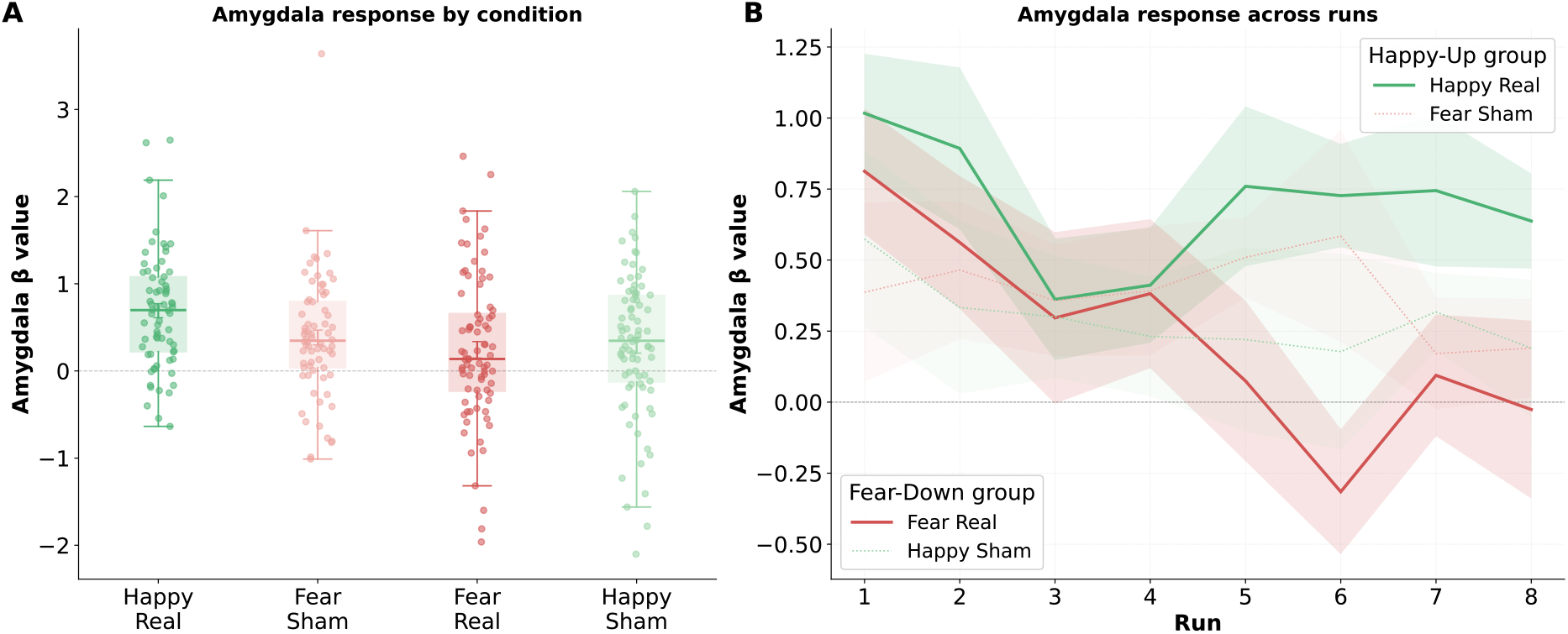
Amygdala regulation performance and changes across neurofeedback training. (A) Distribution of amygdala responses across the four neurofeedback conditions. Amygdala activity was higher during Happy Real than all the other three conditions; the Fear Real versus Fear Sham comparison was in the predicted direction but did not reach significance. (B) Mean bilateral amygdala activity across the eight neurofeedback runs for the Happy Real, Fear Sham, Happy Sham, and Fear Real conditions. Fear downregulation became progressively stronger across training, reflected by decreasing amygdala activity during Fear Real, whereas Happy Real activity was stable across runs.

However, the condition-specific slope analysis provided evidence of significant improvement in the Fear Real condition (Fig. 4B). Amygdala activity decreased significantly across Fear Real runs, b = −0.120, p < .001, consistent with progressive improvement in downregulation. This decrease was significantly steeper than during the Fear Sham condition, b = −0.086, SE = 0.042, t(270.0) = −2.06, p = .020, the Happy Sham condition, b = −0.084, SE = 0.040, t(269.4) = −2.12, p = .017, and the Happy Real condition, b = −0.090, SE = 0.042, t(270.0) = −2.16, p = .016. In contrast, amygdala activity did not increase significantly across Happy Real runs, b = −0.029, p = .827. No significant run-related changes were observed in either sham condition.

#### 2b. Transfer of amygdala regulation

There was no evidence that amygdala responses transferred beyond the neurofeedback training context. For the Fear − Neutral contrast, amygdala activity did not change significantly from baseline to the post-training transfer assessment, b = 0.084, SE = 0.245, p = .733, 95% CI [−0.413, 0.581], or from baseline to follow-up, b = −0.011, SE = 0.245, p = .964, 95% CI [−0.508, 0.486]. The corresponding time × group interactions were also nonsignificant at post-training, b = −0.390, p = .432, and follow-up, b = −0.721, p = .150.

Overall, the No-Feedback Transfer Task provided no evidence of lasting changes in amygdala activity or differential transfer effects between the Happy-up and Fear-down groups. Plots can be found in Supplementary Fig. S1.

### 3. Emotional perception and subjective performance

#### 3a. Emotional perception and arousal

For neutral faces, valence ratings increased significantly from pre-to post-training, b = 4.75, SE = 1.09, p < .001, 95% CI [2.55, 6.96], and remained elevated at follow-up, b = 4.90, SE = 1.11, p < .001, 95% CI [2.65, 7.14] (Fig. 5A). Happy-face valence ratings also increased significantly at post-training, b = 3.56, SE = 1.11, p = .003, 95% CI [1.31, 5.81], and follow-up, b = 2.92, SE = 1.13, p = .014, 95% CI [0.63, 5.22] (Fig. 5E). No significant changes were observed for the valence ratings of fearful faces (Fig. 5C).

**Fig. 5.**
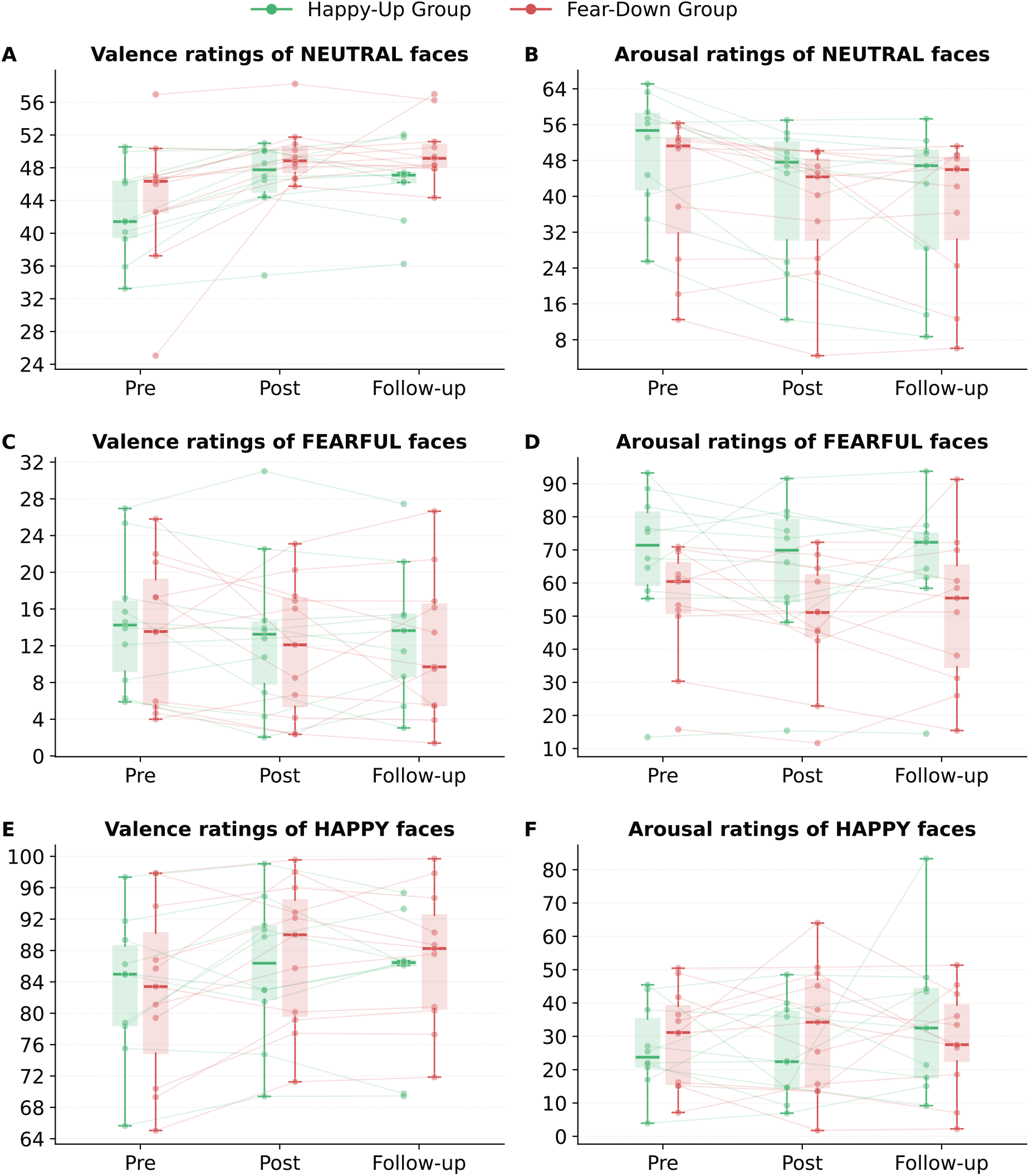
Changes in emotional face ratings (0–100 scale) across the intervention. Valence (or affective perception) and arousal ratings are shown at pre-training, post-training, and follow-up separately for the Happy-up and Fear-down groups. Panels show (A) valence ratings of neutral faces, (B) arousal ratings of neutral faces, (C) valence ratings of fearful faces, (D) arousal ratings of fearful faces, (E) valence ratings of happy faces, and (F) arousal ratings of happy faces. Valence ratings increased for neutral and happy faces following training, and arousal ratings decreased for neutral faces, whereas no significant changes were observed for fearful-face valence ratings or for fearful- and happy-face arousal ratings. No significant time × group interactions were observed.

Arousal ratings for neutral faces decreased significantly at post-training, b = −6.59, SE = 2.12, p = .004, 95% CI [−10.87, −2.30], and follow-up, b = −8.77, SE = 2.16, p < .001, 95% CI [−13.15, −4.39] (Fig. 5B), whereas arousal ratings did not change significantly for fearful (Fig. 5D) or happy faces (Fig. 5F). None of the time × group interactions were significant for either valence or arousal, indicating no evidence that changes differed between the Happy-up and Fear-down groups.

Overall, participants rated neutral and happy faces as more positive and neutral faces as less arousing following training, whereas responses to fearful faces remained unchanged.

#### 3b. Subjective neurofeedback performance

Across all the neurofeedback runs, participants rated their regulation performance significantly higher in the real-feedback conditions than in the sham-feedback conditions, b = 0.31, SE = 0.15, t(309) = 2.01, one-tailed p = .023 (Fig. 6A). Ratings were also higher in the Happy than in the Fear conditions, b = 0.62, SE = 0.15, t(309) = 4.00, p < .001.

**Fig. 6.**
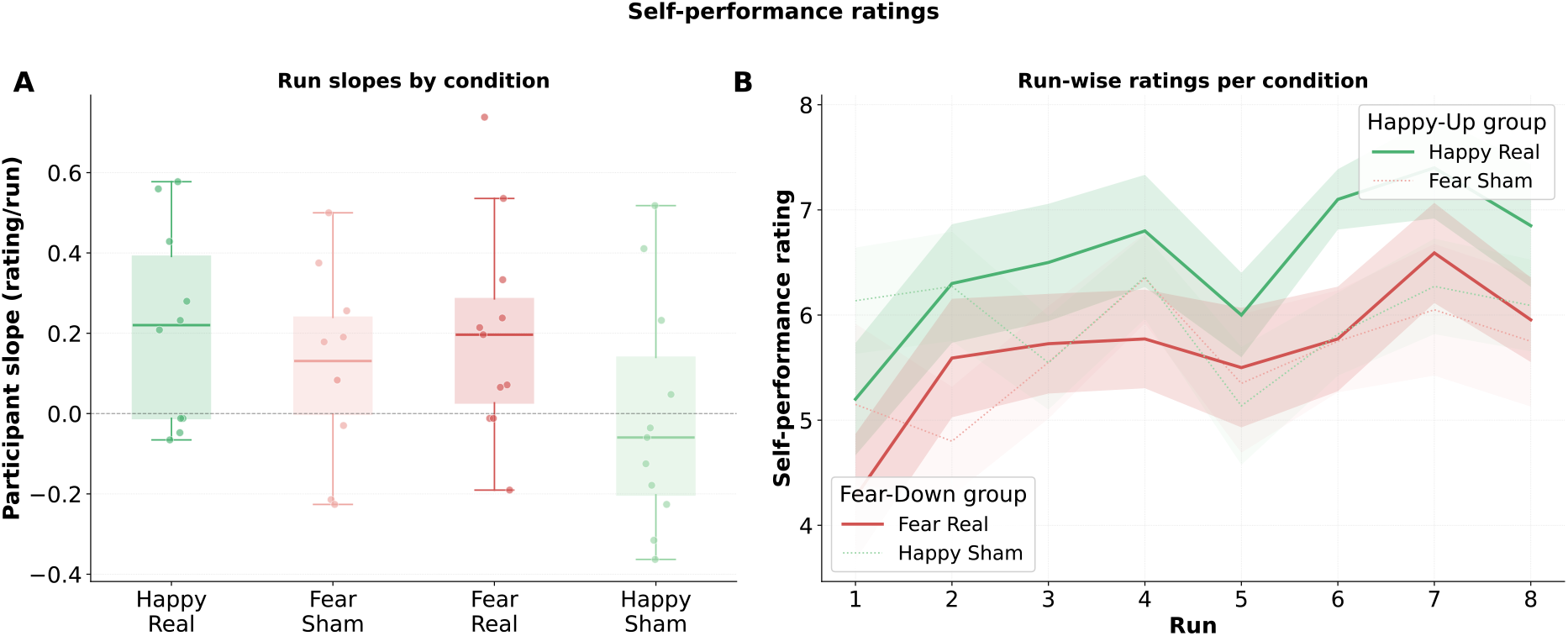
Subjective neurofeedback regulation performance across training. Participants rated their perceived regulation success after each neurofeedback run. Ratings are shown across the eight runs for the Happy Real, Fear Sham, Happy Sham, and Fear Real conditions. (A) Perceived regulation performance was higher overall during real than sham feedback and (B) increased significantly across runs in both real-feedback conditions, whereas no significant linear changes were observed in the sham conditions.

Subjective performance ratings increased across runs, b = 0.131, SE = 0.034, t(309) = 3.90, p < .001, and this increase was significantly steeper for real than for sham feedback, b = 0.151, SE = 0.067, t(309) = 2.25, p = .025. Ratings increased significantly in both real-feedback conditions: in the Happy Real condition, b = 0.215, SE = 0.069, p = .002, and in the Fear Real condition, b = 0.198, SE = 0.066, p = .003. No significant linear changes were observed in either sham condition (Fig. 6B). The remaining interactions were not significant.

Overall, participants perceived their regulation performance as better during real than sham feedback and reported increasing perceived success across training in both real-feedback conditions.

### 4. Brain–clinical associations

In the Fear-down group, mean amygdala activity during the Fear Real condition was negatively associated with change in MADRS scores, ρ = −.857, 95% CI [−.966, −.495], p = .002, and STAI-S scores, ρ = −.722, 95% CI [−.929, −.170], p = .018, and positively associated with change in MDBF calmness scores, ρ = .634, 95% CI [.007, .903], p = .049, after controlling for pre-training scores (Fig. S2). However, only the association with MADRS change survived Holm–Bonferroni correction, p_adj_ = .014. No significant associations were observed for HAM-D, STAI-T, SHAPS, sense of control, good mood, or wakefulness.

In the Happy-up group, mean amygdala activity during the Happy Real condition was negatively associated with change in HAM-D scores, ρ = −.824, 95% CI [−.962, −.353], p = .006, but this association did not survive correction, p_adj_ = .057. None of the other correlations reached significance. Plots can be found in Supplementary Fig. S2.

In the sham conditions, no associations survived Holm–Bonferroni correction.

## Discussion

This study investigated whether a naturalistic face-based neurofeedback paradigm could engage distinct affective processes involved in MDD and produce symptom-specific clinical changes. The paradigm, previously tested in healthy participants^23^, was designed as an adaptive, self-reinforcing closed-loop system in which the feedback was embedded in the emotional stimulus itself. In the Happy condition, amygdala upregulation shifted the face toward a happier expression, whereas in the Fear condition, amygdala downregulation shifted the face toward a less fearful, more neutral expression. These changes in facial expression were in turn intended to evoke amygdala activity towards the targeted regulatory direction. Participants in the Happy-up and Fear-down groups trained amygdala upregulation and downregulation, respectively, to target different affective processes. Specifically, upregulation in response to happy faces aimed to enhance responsiveness to positive emotional information, a process relevant to anhedonia, whereas downregulation in response to fearful faces aimed to attenuate responsiveness to negative emotional information, a process relevant to anxiety.

At the neural level, both training conditions showed evidence of amygdala regulation in the predicted direction, although their temporal profiles differed. Amygdala upregulation in the Happy condition was characterized by higher amygdala activity during real feedback compared with both sham feedback conditions and the Fear Real condition across training, without a systematic increase across runs. In the Fear condition, the overall real-versus-sham difference was not significant across the full training period, but downregulation became progressively stronger over successive runs, with amygdala activity decreasing significantly during real feedback and more steeply than during the two sham conditions, as well as the Happy Real condition. Thus, both conditions showed amygdala regulation in the intended direction through different temporal trajectories. Upregulation in the Happy condition appeared to be present from early training and was subsequently maintained, whereas downregulation in the Fear condition improved progressively with practice.

The same pattern was observed in our previous study in healthy participants^23^, in which downregulation in the Fear condition improved across runs, whereas upregulation in the Happy condition showed no significant run-wise change. Its recurrence in patients with MDD suggests that this pattern is not specific to the disorder. It remains unclear, however, whether these different training trajectories reflect differences in the difficulty of amygdala upregulation versus downregulation or the influence of emotional valence, since upregulation was always trained with happy faces and downregulation with fearful faces. With respect to regulation direction, up- and downregulation of emotion recruit partly distinct neural systems, both during amygdala neurofeedback^68^ and without neurofeedback^69^, with downregulation engaging dorsolateral prefrontal control regions more strongly^70^. With respect to emotional valence, regulating highly salient negative emotional information may place greater demands on cognitive control and strategy selection^71^, while cognitive-control training has been shown to facilitate subsequent regulation of negative emotion^72^. Together, these findings suggest that Fear downregulation may have been more demanding and therefore required more practice to achieve effective regulation.

Embedding the feedback signal into the emotional stimulus is the central design feature of the paradigm. Previous research has shown that the format of neurofeedback can influence neural regulation, with socially meaningful feedback, such as a smiling avatar, associated with greater activation of the targeted brain region than conventional bar feedback^73^. Beyond the social relevance of the feedback, the present paradigm integrated the neurofeedback signal directly into the emotional stimulus being regulated. This approach is consistent with the process-based neurofeedback framework proposed by Lubianiker et al.^74^, which advocates linking neural targets and feedback environments to clinically relevant psychological processes. In the present study, participants showed amygdala regulation during training and reported greater perceived regulation success during real than sham feedback. Although these findings support the use of dynamic emotional faces as a neurofeedback interface in patients with MDD, the extent to which the changing expressions themselves facilitated regulation remains unknown.

Despite the observed neural regulation, the intervention did not produce the hypothesized symptom-specific clinical effects. Depression severity, anhedonia, and anxiety improved over the course of the study, but these changes did not differ significantly between the Happy-up and Fear-down groups. This pattern can be considered in light of clinical research showing that interventions targeting distinct affective processes may produce overlapping benefits. In randomized trials comparing positive-affect treatment targeting reward sensitivity with negative-affect treatment targeting threat sensitivity, Craske et al.^75,76^ found that enhancing positive affect improved not only positive-affect outcomes but also depression and anxiety symptoms. These findings suggest that interventions targeting positive and negative affect do not necessarily produce improvements restricted to their respective symptom dimensions. However, the absence of between-group differences in the present study does not establish that the two neurofeedback approaches have equivalent clinical effects.

The reductions in depression severity were notable given the brief training period, with mean decreases of 4.04 points (27.1%) on the HAM-D and 6.52 points (31.7%) on the MADRS. These changes were observed after only two neurofeedback training sessions on consecutive days, with post-training assessments conducted three days after baseline. The magnitude of the reductions was comparable to conventional definitions of partial clinical response^77^, although these definitions apply to individual patients rather than changes in group means. The MADRS reduction also exceeded published estimates of minimal important differences^78^, although these estimates were developed to evaluate antidepressant treatment effects rather than uncontrolled pre–post changes. The limited training duration may have been insufficient for the distinct trajectories of neural regulation to translate into differential clinical outcomes. Future studies with extended training protocols could examine whether additional sessions lead to greater clinical improvements or more selective effects on depressive symptoms.

Whether the observed clinical improvements were specifically attributable to neurofeedback remains unclear. Previous randomized controlled trials in MDD have reported greater clinical improvements following amygdala neurofeedback compared with control feedback^26^, whereas other studies found comparable improvements following neurofeedback targeting emotion-related and control brain regions^25^. In the latter study, clinical improvement was associated with increased self-efficacy, suggesting that the experience of successful self-regulation may contribute to therapeutic benefits independently of the specific neural target. Comparable clinical improvements between active and control neurofeedback conditions have also been reported in alcohol dependence and posttraumatic stress disorder^79,80^. This literature suggests that shared psychological processes, such as perceived control, expectancy, and emotion-regulation practice, may contribute to clinical improvement. In the present study, all participants received real feedback in one condition and sham feedback in the other, and group assignment alternated rather than being randomized. Without a sham-only clinical control group, alternative explanations for improvement over time, including regression to the mean, cannot be excluded. Exploratory analyses also revealed associations between amygdala activity during neurofeedback and clinical improvement. In both the Happy-up and Fear-down groups, higher mean amygdala activity during real feedback was associated with greater reductions in depression severity. Given the small group-specific samples and the exploratory nature of these analyses, these findings should be interpreted cautiously despite the large observed correlations.

Changes in facial-affect perception provide an additional indication that participants’ emotional processing changed over the course of the intervention. Across groups, neutral and happy faces were rated more positively following training, and these effects persisted at follow-up. This pattern is potentially relevant to MDD, in which neutral and ambiguous facial expressions are often interpreted with a negative bias^41,43^. However, because these effects did not differ between the Happy-up and Fear-down groups, they cannot be attributed to the condition-specific neurofeedback mechanisms. They may instead reflect repeated engagement with emotional facial expressions, emotion-regulation practice, or broader changes occurring during the intervention. These behavioural changes are therefore informative, but should be distinguished from the evidence for condition-specific neural regulation.

### Limitations

Several limitations should be considered. The bilateral amygdala target was defined anatomically despite substantial functional heterogeneity within the amygdala^81,82^. Individualized or functionally defined targets may offer greater neurofeedback specificity, although whether such approaches consistently improve neurofeedback learning remains unclear^50^. Regarding clinical outcomes, relatively mild baseline anhedonia may have limited the sensitivity to detect improvements following Happy-up training. Although the Happy-up and Fear-down conditions were designed to target processes relevant to anhedonia and anxiety, respectively, participants were assigned to training independently of their individual symptom profiles. The study therefore evaluated symptom-targeted neurofeedback without investigating personalized treatment allocation. Future studies could examine whether matching neurofeedback interventions to individual symptom profiles improves clinical outcomes.

Small sample sizes represent a broader challenge in rtfMRI-NF research^83,84^, where extensive repeated scanning provides rich within-subject data but cannot fully compensate for limited statistical power at the participant level. Here, each patient underwent five scanning sessions, enabling detailed characterization of neural regulation despite the relatively small sample.

Evidence for transfer beyond the neurofeedback context was also inconclusive. Regulation effects observed during training were not reproduced during the No-Feedback Transfer Task. However, this absence of a significant transfer effect should be interpreted cautiously, as successful regulation during feedback does not necessarily generalize to conditions in which feedback is absent^50^. Moreover, the transfer assessment consisted of a single, relatively short fMRI run, which may have limited sensitivity to detect group-by-time effects across repeated assessments^85^. The present data therefore do not provide strong evidence either for or against generalization of the acquired regulation skills beyond the training context.

## Conclusion

Patients with MDD demonstrated amygdala upregulation and downregulation using dynamic facial expressions as the feedback signal, with each regulation direction trained in a separate group. Upregulation in the Happy condition was present from early training, whereas downregulation in the Fear condition improved progressively across runs. Depression severity, anhedonia, and anxiety decreased following training, but these improvements did not differ between the Happy-up and Fear-down groups. More broadly, the results support dynamic social-emotional stimuli as a feasible means of embedding the neurofeedback signal within the affective context of regulation. Future studies could test whether symptom- or biomarker-guided treatment allocation can translate differential neural regulation into more selective clinical benefit.

## Data Availability

The data supporting the findings of this study are available from the corresponding author upon request, subject to approval through an institutional data-sharing agreement.

https://github.com/Pichlore/clamy-mdd

## Acknowledgements

We thank Gianluca Macauda for the technical assistance as well as Zachary Roman and Filip Melinscak for data analysis advice.

Use of AI tools: Claude (Anthropic) and Copilot were used to optimize the analysis code. ChatGPT and Claude were used to edit the manuscript text for language and clarity. All content was reviewed and verified by the authors, who take full responsibility for the integrity and accuracy of the work.

## Author contributions

AW, RS, FS, AB, AH, DW, CL and BK contributed to the experimental design and the conceptualization of the study. ML was responsible for patient recruitment. JA and KM assisted with data collection. PS provided technical support. MZ contributed to the data analysis. AW coordinated data collection. AW and CL performed the data analysis and drafted the manuscript. FS acquired funding and supervised the project. All authors critically revised the manuscript and approved the final version.

## Funding

This study was supported by the Swiss National Science Foundation (grants BSSG10_155915, 100014_178841 and 32003B_166566), the Foundation for Research in Science and the Humanities at the University of Zurich (STWF-17-012), the Baugarten Stiftung, and the Austrian Research Promotion Agency (FFG; Project Nos. FO999927913). AH was supported by the Fondation Adrian et Simone Frutiger and the Swiss National Science Foundation (project number 233167).

## Competing interests

The authors declare no competing interests.

## Code availability

The neurofeedback implementation was based on OpenNFT (https://github.com/OpenNFT/OpenNFT). The analysis code is available at https://github.com/Pichlore/clamy-mdd.

## Ethics approval and consent to participate

The study was approved by the Kantonale Ethikkommission Zürich and was conducted in accordance with the Declaration of Helsinki. All participants provided written informed consent prior to participation.

## Notes

### Competing Interest Statement

The authors have declared no competing interest.

### Clinical Trial

ISRCTN71745131

### Clinical Protocols

https://doi.org/10.1186/ISRCTN71745131

### Author Declarations

Ethics Committee of the Canton of Zurich gave ethical approval for this work

## References

1. Athira, K. V. et al. An Overview of the Heterogeneity of Major Depressive Disorder: Current Knowledge and Future Prospective. Curr Neuropharmacol 18, 168–187 (2020).

2. Dold, M. et al. Major Depression and the Degree of Suicidality: Results of the European Group for the Study of Resistant Depression (GSRD). International Journal of Neuropsychopharmacology 21, 539–549 (2018).

3. Fried, E. I. & Nesse, R. M. Depression is not a consistent syndrome: An investigation of unique symptom patterns in the STAR*D study. Journal of Affective Disorders 172, 96–102 (2015).

4. Santomauro, D. F. et al. Global prevalence and burden of depressive and anxiety disorders in 204 countries and territories in 2020 due to the COVID-19 pandemic. The Lancet 398, 1700–1712 (2021).

5. Kessler, R. C. & Bromet, E. J. The Epidemiology of Depression Across Cultures. Annu. Rev. Public Health 34, 119–138 (2013).

6. Beck, A. T. The Evolution of the Cognitive Model of Depression and Its Neurobiological Correlates. AJP 165, 969–977 (2008).

7. Buch, A. M. & Liston, C. Dissecting diagnostic heterogeneity in depression by integrating neuroimaging and genetics. Neuropsychopharmacol. 46, 156–175 (2021).

8. Insel, T. et al. Research Domain Criteria (RDoC): Toward a New Classification Framework for Research on Mental Disorders. AJP 167, 748–751 (2010).

9. Csukly, G. et al. What is the relationship between the recognition of emotions and core beliefs: Associations between the recognition of emotions in facial expressions and the maladaptive schemas in depressed patients. Journal of Behavior Therapy and Experimental Psychiatry 42, 129–137 (2011).

10. McMakin, D. L. et al. Anhedonia Predicts Poorer Recovery Among Youth With Selective Serotonin Reuptake Inhibitor Treatment–Resistant Depression. Journal of the American Academy of Child & Adolescent Psychiatry 51, 404–411 (2012).

11. Disner, S. G., Beevers, C. G., Haigh, E. A. P. & Beck, A. T. Neural mechanisms of the cognitive model of depression. Nat Rev Neurosci 12, 467–477 (2011).

12. Roiser, J. P., Elliott, R. & Sahakian, B. J. Cognitive Mechanisms of Treatment in Depression. Neuropsychopharmacol 37, 117–136 (2012).

13. Gothard, K. M. Multidimensional processing in the amygdala. Nat Rev Neurosci 21, 565–575 (2020).

14. Groenewold, N. A., Opmeer, E. M., De Jonge, P., Aleman, A. & Costafreda, S. G. Emotional valence modulates brain functional abnormalities in depression: Evidence from a meta-analysis of fMRI studies. Neuroscience & Biobehavioral Reviews 37, 152–163 (2013).

15. Siegle, G. J., Thompson, W., Carter, C. S., Steinhauer, S. R. & Thase, M. E. Increased Amygdala and Decreased Dorsolateral Prefrontal BOLD Responses in Unipolar Depression: Related and Independent Features. Biological Psychiatry 61, 198–209 (2007).

16. Suslow, T. et al. Automatic Mood-Congruent Amygdala Responses to Masked Facial Expressions in Major Depression. Biological Psychiatry 67, 155–160 (2010).

17. Victor, T. A., Furey, M. L., Fromm, S. J., Öhman, A. & Drevets, W. C. Relationship Between Amygdala Responses to Masked Faces and Mood State and Treatment in Major Depressive Disorder. Arch Gen Psychiatry 67, 1128 (2010).

18. Hawkinson, J. E. et al. Quantification of Adverse Events Associated with Functional MRI Scanning and with Real-Time fMRI-Based Training. *Int.J*. Behav. Med. 19, 372–381 (2012).

19. Sulzer, J. et al. Real-time fMRI neurofeedback: Progress and challenges. NeuroImage 76, 386– 399 (2013).

20. Weiskopf, N. et al. Real-time functional magnetic resonance imaging: methods and applications. Magnetic Resonance Imaging 25, 989–1003 (2007).

21. Sitaram, R. et al. Closed-loop brain training: the science of neurofeedback. Nat Rev Neurosci 18, 86–100 (2017).

22. Herwig, U. et al. Training emotion regulation through real-time fMRI neurofeedback of amygdala activity. NeuroImage 184, 687–696 (2019).

23. Watve, A. et al. Facing emotions: real-time fMRI-based neurofeedback using dynamic emotional faces to modulate amygdala activity. Front. Neurosci. 17, 1286665 (2024).

24. Linden, D. E. J. et al. Real-Time Self-Regulation of Emotion Networks in Patients with Depression. PLoS ONE 7, e38115 (2012).

25. Mehler, D. M. A. et al. Targeting the affective brain—a randomized controlled trial of real-time fMRI neurofeedback in patients with depression. Neuropsychopharmacol 43, 2578–2585 (2018).

26. Young, K. D. et al. Randomized Clinical Trial of Real-Time fMRI Amygdala Neurofeedback for Major Depressive Disorder: Effects on Symptoms and Autobiographical Memory Recall. AJP 174, 748–755 (2017).

27. Young, K. D. et al. Real-Time fMRI Neurofeedback Training of Amygdala Activity in Patients with Major Depressive Disorder. PLoS ONE 9, e88785 (2014).

28. Zotev, V., Mayeli, A., Misaki, M. & Bodurka, J. Emotion self-regulation training in major depressive disorder using simultaneous real-time fMRI and EEG neurofeedback. NeuroImage: Clinical 27, 102331 (2020).

29. Tsuchiyagaito, A. et al. Real-Time fMRI Functional Connectivity Neurofeedback Reducing Repetitive Negative Thinking in Depression: A Double-Blind, Randomized, Sham-Controlled Proof-of-Concept Trial. Psychother Psychosom 92, 87–100 (2023).

30. Trambaiolli, L. R., Kohl, S. H., Linden, D. E. J. & Mehler, D. M. A. Neurofeedback training in major depressive disorder: A systematic review of clinical efficacy, study quality and reporting practices. Neuroscience & Biobehavioral Reviews 125, 33–56 (2021).

31. Morris, L. S. et al. Targeting the dopaminergic midbrain with precision 7-Tesla biofeedback training in depression: A proof-of-principle randomized controlled trial. Mol Psychiatry 10.1038/s41380-026-03911-x (2026)

32. Young, K. D. et al. Real-Time Functional Magnetic Resonance Imaging Amygdala Neurofeedback Changes Positive Information Processing in Major Depressive Disorder. Biological Psychiatry 82, 578–586 (2017).

33. Paret, C. et al. Down-regulation of amygdala activation with real-time fMRI neurofeedback in a healthy female sample. Front. Behav. Neurosci. 8, (2014).

34. Paret, C. et al. fMRI neurofeedback of amygdala response to aversive stimuli enhances prefrontal–limbic brain connectivity. NeuroImage 125, 182–188 (2016).

35. Brühl, A. B. et al. Real-time Neurofeedback Using Functional MRI Could Improve Down-Regulation of Amygdala Activity During Emotional Stimulation: A Proof-of-Concept Study. Brain Topogr 27, 138–148 (2014).

36. Compère, L. et al. Enhanced efficacy of CBT following augmentation with amygdala rtfMRI neurofeedback in depression. Journal of Affective Disorders 339, 495–501 (2023).

37. Compère, L. et al. Amygdala real-time fMRI neurofeedback upregulation in treatment resistant depression: Proof of concept and dose determination. Behaviour Research and Therapy 176, 104523 (2024).

38. Sarkheil, P. et al. fMRI feedback enhances emotion regulation as evidenced by a reduced amygdala response. Behavioural Brain Research 281, 326–332 (2015).

39. Kupferberg, A., Bicks, L. & Hasler, G. Social functioning in major depressive disorder. Neuroscience & Biobehavioral Reviews 69, 313–332 (2016).

40. Kupferberg, A. & Hasler, G. The social cost of depression: Investigating the impact of impaired social emotion regulation, social cognition, and interpersonal behavior on social functioning. Journal of Affective Disorders Reports 14, 100631 (2023).

41. Weightman, M. J., Air, T. M. & Baune, B. T. A Review of the Role of Social Cognition in Major Depressive Disorder. Front. Psychiatry 5, (2014).

42. Monferrer, M. et al. Facial emotion recognition in patients with depression compared to healthy controls when using human avatars. Sci Rep 13, 6007 (2023).

43. Münkler, P., Rothkirch, M., Dalati, Y., Schmack, K. & Sterzer, P. Biased Recognition of Facial Affect in Patients with Major Depressive Disorder Reflects Clinical State. PLoS ONE 10, e0129863 (2015).

44. Bourke, C., Douglas, K. & Porter, R. Processing of Facial Emotion Expression in Major Depression: A Review. Aust N Z J Psychiatry 44, 681–696 (2010).

45. Penton-Voak, I. S., Bate, H., Lewis, G. & Munafò, M. R. Effects of emotion perception training on mood in undergraduate students: randomised controlled trial. Br J Psychiatry 201, 71–72 (2012).

46. Tranter, R. et al. The effect of serotonergic and noradrenergic antidepressants on face emotion processing in depressed patients. Journal of Affective Disorders 118, 87–93 (2009).

47. Quevedo, K. et al. Neurofeedback and neuroplasticity of visual self-processing in depressed and healthy adolescents: A preliminary study. Developmental Cognitive Neuroscience 40, 100707 (2019).

48. Hartwell, K. J. et al. Individualized real-time fMRI neurofeedback to attenuate craving in nicotine-dependent smokers. J Psychiatry Neurosci 41, 48–55 (2016).

49. Ihssen, N., Sokunbi, M. O., Lawrence, A. D., Lawrence, N. S. & Linden, D. E. J. Neurofeedback of visual food cue reactivity: a potential avenue to alter incentive sensitization and craving. Brain Imaging and Behavior 11, 915–924 (2017).

50. Paret, C. et al. Current progress in real-time functional magnetic resonance-based neurofeedback: Methodological challenges and achievements. NeuroImage 202, 116107 (2019).

51. Krumhuber, E. G., Skora, L. I., Hill, H. C. H. & Lander, K. The role of facial movements in emotion recognition. Nat Rev Psychol 2, 283–296 (2023).

52. Sheehan, D. V. et al. The Mini-International Neuropsychiatric Interview (M.I.N.I.): the development and validation of a structured diagnostic psychiatric interview for DSM-IV and ICD-10. J Clin Psychiatry **59 Suppl 20**, 22-33;quiz 34-57 (1998).

53. Ros, T. et al. Consensus on the reporting and experimental design of clinical and cognitive-behavioural neurofeedback studies (CRED-nf checklist). Brain 143, 1674–1685 (2020).

54. Tyszka, J. M. & Pauli, W. M. In vivo delineation of subdivisions of the human amygdaloid complex in a high-resolution group template. Human Brain Mapping 37, 3979–3998 (2016).

55. Langner, O. et al. Presentation and validation of the Radboud Faces Database. Cognition and Emotion 24, 1377–1388 (2010).

56. Willinger, D. et al. Valence-Dependent Coupling of Prefrontal-Amygdala Effective Connectivity during Facial Affect Processing. eNeuro 6, ENEURO.0079-19.2019 (2019).

57. Koush, Y. et al. OpenNFT: An open-source Python/Matlab framework for real-time fMRI neurofeedback training based on activity, connectivity and multivariate pattern analysis. NeuroImage 156, 489–503 (2017).

58. Sorger, B., Scharnowski, F., Linden, D. E. J., Hampson, M. & Young, K. D. Control freaks: Towards optimal selection of control conditions for fMRI neurofeedback studies. NeuroImage 186, 256–265 (2019).

59. Peirce, J. et al. PsychoPy2: Experiments in behavior made easy. Behav Res 51, 195–203 (2019).

60. Hamilton, M. A rating scale for depression. *Journal of Neurology*, Neurosurgery & Psychiatry 23, 56–62 (1960).

61. Montgomery, S. A. & Åsberg, M. A New Depression Scale Designed to be Sensitive to Change. Br J Psychiatry 134, 382–389 (1979).

62. Bieling, P. J., Antony, M. M. & Swinson, R. P. The State--Trait Anxiety Inventory, Trait version: structure and content re-examined. Behaviour Research and Therapy 36, 777–788 (1998).

63. Franz, M. et al. Deutsche Version der Snaith-Hamilton-Pleasure-Scale (SHAPS-D). Fortschr Neurol Psychiatr 66, 407–413 (1998).

64. Steyer, R., Schwenkmezger, P., Notz, P. & Eid, M. Mehrdimensionalen Befindlichkeitsfragebogen. 10.1037/t12446-000 (2017).

65. Krampen, G. Fragebogen zu Kompetenz-und Kontrollüberzeugungen (FKK) [Questionnaire on competence and control beliefs] [Manual]. (Hogrefe, Göttingen, 1991).

66. Kopel, R. et al. No time for drifting: Comparing performance and applicability of signal detrending algorithms for real-time fMRI. NeuroImage 191, 421–429 (2019).

67. Haugg, A. et al. Predictors of real-time fMRI neurofeedback performance and improvement – A machine learning mega-analysis. NeuroImage 237, 118207 (2021).

68. Paret, C. et al. Monitoring and control of amygdala neurofeedback involves distributed information processing in the human brain. Human Brain Mapping 39, 3018–3031 (2018).

69. Min, J. et al. Emotion Downregulation Targets Interoceptive Brain Regions While Emotion Upregulation Targets Other Affective Brain Regions. J. Neurosci. 42, 2973–2985 (2022).

70. Sokołowski, A., Morawetz, C., Folkierska-Żukowska, M. & Łukasz Dragan, W. Brain activation during cognitive reappraisal depending on regulation goals and stimulus valence. Social Cognitive and Affective Neuroscience 17, 559–570 (2022).

71. Sheppes, G., Scheibe, S., Suri, G. & Gross, J. J. Emotion-Regulation Choice. Psychol Sci 22, 1391–1396 (2011).

72. Cohen, N. & Mor, N. Enhancing Reappraisal by Linking Cognitive Control and Emotion. Clinical Psychological Science 6, 155–163 (2018).

73. Mathiak, K. A. et al. Social reward improves the voluntary control over localized brain activity in fMRI-based neurofeedback training. Front. Behav. Neurosci. 9, (2015).

74. Lubianiker, N. et al. Process-based framework for precise neuromodulation. Nat Hum Behav 3, 436–445 (2019).

75. Craske, M. G., Meuret, A. E., Echiverri-Cohen, A., Rosenfield, D. & Ritz, T. Positive affect treatment targets reward sensitivity: A randomized controlled trial. Journal of Consulting and Clinical Psychology 91, 350–366 (2023).

76. Craske, M. G. et al. Positive affect treatment for depression and anxiety: A randomized clinical trial for a core feature of anhedonia. Journal of Consulting and Clinical Psychology 87, 457–471 (2019).

77. Lam, R. W. et al. Canadian Network for Mood and Anxiety Treatments (CANMAT) 2023 Update on Clinical Guidelines for Management of Major Depressive Disorder in Adults: Réseau canadien pour les traitements de l’humeur et de l’anxiété (CANMAT) 2023 : Mise à jour des lignes directrices cliniques pour la prise en charge du trouble dépressif majeur chez les adultes. Can J Psychiatry 69, 641–687 (2024).

78. Hengartner, M. P. & Plöderl, M. Estimates of the minimal important difference to evaluate the clinical significance of antidepressants in the acute treatment of moderate-to-severe depression. BMJ EBM 27, 69–73 (2022).

79. Karch, S. et al. Functional and clinical outcomes of FMRI-based neurofeedback training in patients with alcohol dependence: a pilot study. Eur Arch Psychiatry Clin Neurosci 272, 557–569 (2022).

80. Zhao, Z. et al. Amygdala downregulation training using fMRI neurofeedback in post-traumatic stress disorder: a randomized, double-blind trial. Transl Psychiatry 13, 177 (2023).

81. Sladky, R., Kargl, D., Haubensak, W. & Lamm, C. An active inference perspective for the amygdala complex. Trends in Cognitive Sciences 28, 223–236 (2024).

82. Sladky, R., Riva, F., Rosenberger, L. A., Van Honk, J. & Lamm, C. Basolateral and central amygdala orchestrate how we learn whom to trust. Commun Biol 4, 1329 (2021).

83. Fede, S. J., Dean, S. F., Manuweera, T. & Momenan, R. A Guide to Literature Informed Decisions in the Design of Real Time fMRI Neurofeedback Studies: A Systematic Review. Front. Hum. Neurosci. 14, 60 (2020).

84. Tursic, A., Eck, J., Lührs, M., Linden, D. E. J. & Goebel, R. A systematic review of fMRI neurofeedback reporting and effects in clinical populations. NeuroImage: Clinical 28, 102496 (2020).

85. Friedman, L. et al. Test–retest and between-site reliability in a multicenter fMRI study. Human Brain Mapping 29, 958–972 (2008).

